# Overall and cause-specific hospitalisation and death after COVID-19 hospitalisation in England: cohort study in OpenSAFELY using linked primary care, secondary care and death registration data

**DOI:** 10.1101/2021.07.16.21260628

**Authors:** Krishnan Bhaskaran, Christopher T Rentsch, George Hickman, William J Hulme, Anna Schultze, Helen J Curtis, Kevin Wing, Charlotte Warren-Gash, Laurie Tomlinson, Chris J Bates, Rohini Mathur, Brian MacKenna, Viyaasan Mahalingasivam, Angel Wong, Alex J Walker, Caroline E Morton, Daniel Grint, Amir Mehrkar, Rosalind M Eggo, Peter Inglesby, Ian J Douglas, Helen I McDonald, Jonathan Cockburn, Elizabeth J Williamson, David Evans, John Parry, Frank Hester, Sam Harper, Stephen JW Evans, Sebastian Bacon, Liam Smeeth, Ben Goldacre

## Abstract

**Background:** There is concern about medium to long-term adverse outcomes following acute COVID-19, but little relevant evidence exists. We aimed to investigate whether risks of hospital admission and death, overall and by specific cause, are raised following discharge from a COVID-19 hospitalisation.

**Methods and Findings:** Working on behalf of NHS-England, we used linked primary care and hospital data in OpenSAFELY to compare risks of hospital admission and death, overall and by specific cause, between people discharged from COVID-19 hospitalisation (February-December 2020), and (i) demographically-matched controls from the 2019 general population; (ii) people discharged from influenza hospitalisation in 2017-19. We used Cox regression adjusted for personal and clinical characteristics.

24,673 post-discharge COVID-19 patients, 123,362 general population controls, and 16,058 influenza controls were followed for ≤315 days. Overall risk of hospitalisation or death (30968 events) was higher in the COVID-19 group than general population controls (adjusted-HR 2.23, 2.14-2.31) but similar to the influenza group (adjusted-HR 0.94, 0.91-0.98). All-cause mortality (7439 events) was highest in the COVID-19 group (adjusted-HR 4.97, 4.58-5.40 vs general population controls and 1.73, 1.60-1.87 vs influenza controls). Risks for cause-specific outcomes were higher in COVID-19 survivors than general population controls, and largely comparable between COVID-19 and influenza patients. However, COVID-19 patients were more likely than influenza patients to be readmitted/die due to their initial infection/other lower respiratory tract infection (adjusted-HR 1.37, 1.22-1.54), and to experience mental health or cognitive-related admission/death (adjusted-HR 1.36, 1.01-2.83); in particular, COVID-19 survivors with pre-existing dementia had higher risk of dementia death. One limitation of our study is that reasons for hospitalisation/death may have been misclassified in some cases due to inconsistent use of codes.

**Conclusions:** People discharged from a COVID-19 hospital admission had markedly higher risks for rehospitalisation and death than the general population, suggesting a substantial extra burden on healthcare. Most risks were similar to those observed after influenza hospitalisations; but COVID-19 patients had higher risks of all-cause mortality, readmissions/death due to the initial infection, and dementia death, highlighting the importance of post-discharge monitoring.

## Introduction

Severe acute respiratory syndrome coronavirus 2 (SARS-CoV-2) emerged in early 2020 and rapidly spread around the world, infecting >140 million people globally.^1^ Acute infection can be asymptomatic or mild,^2^ but a substantial minority of infected people experience severe COVID-19 requiring hospitalisation,^3^ with age being a major risk factor, along with male sex, non-White ethnicity and certain comorbidities.^4-6^ Early in the pandemic, the proportion surviving hospitalisation was around 50-70%,^7^ though improved treatment guidelines and the identification of effective therapies such as dexamethasone helped to improve survival rates.^8,9^ There is now a large and growing population of people who have survived a COVID-19 hospitalisation, but little is known about their longer-term health outcomes.

Emerging evidence suggests that a subset of people infected with SARS-CoV-2 can experience health problems for at least several months after the acute phase of their infection, with fatigue, pain, respiratory and cardiovascular symptoms, and mental health and cognitive disturbances being among the problems frequently described under the term “post-acute COVID-19 syndrome”;^10^ however, epidemiological characterisation of such sequelae remains immature. Small descriptive studies of COVID-19 survivors have been suggestive of high incidence of a range of outcomes including respiratory, cardiovascular, and mental-health related, but firm conclusions are difficult due to lack of comparison groups.^11,12^ There remains limited evidence about post-COVID sequelae across the full range of health outcomes. One recent study of US Department of Veterans Affairs (VA) data examined a wide range of diagnoses, prescriptions and laboratory abnormalities among 30-day survivors of COVID-19, showing excess risks of several health outcomes in the 6 months following infection, compared with the general VA population.^13^ Whether these findings generalise to the entire US population or other settings remains unclear. Another US study limited to people aged <65 years also found excess risks of a range of clinical outcomes ascertained from health insurance data among people with a record of SARS-CoV-2 infection.^14^ A UK study of routinely-collected primary care and hospitalisation data described raised rates of all-cause hospital admission and death among patients discharged following a COVID-19 hospitalisation; the authors also noted raised risks of adverse respiratory and cardiovascular sequalae among the selected outcomes investigated.^15^ Only a general population comparator was used, making it difficult to disentangle risks specific to COVID-19 from those associated with hospitalisation more generally; furthermore, a hospitalised cohort is likely to have been more prone to health problems at the outset than the general population comparator group.

To strengthen the evidence base in this important emerging area, we therefore aimed to investigate the incidence of subsequent hospital admission and death, both overall and from a wide range of specific causes, following a COVID-19 hospitalisation in England. We aimed to compare post-COVID risks with two separate comparison groups: (i) the general population, and (ii) people hospitalised for influenza prior to the current pandemic.

## Methods

### Study design and study population

A cohort study was carried out within the OpenSAFELY platform, which has been described previously.^6^ We used routinely-collected electronic data from primary care practices using TPP SystmOne software, covering approximately 40% of the population in England, linked at the individual patient level to NHS Secondary Uses Service (SUS) data on hospitalisations, and Office of National Statistics death registration data (from 2019 onwards). We selected all individuals discharged between 1^st^ February and 30^th^ December 2020 from a hospitalisation that lasted >1 day and where COVID-19 was coded as the primary diagnosis (based on the International Classification of Diseases (ICD)-10 codes U07.1 “COVID-19 - virus identified” and U07.2 “COVID-19 - virus not identified”) and who were alive and under follow-up one week after discharge (to avoid a focus on hospital transfers and immediate readmissions/deaths). We excluded a small number of people with missing age, sex, or index of multiple deprivation, which are likely to indicate poor data quality. Two comparison groups were also selected: (i) people under follow-up in the general population in 2019, individually matched 5:1 to the COVID-19 group on age (within 3 years), sex, Sustainability and Transformation Plans (STP, a geographical area used as in NHS administration, of which there were 32 in our data), and calendar month (e.g. a patient discharged from a COVID-19 hospitalisation in April 2020 was matched to 5 individuals of the same age, sex and STP who were under follow-up in general practice on 1^st^ April 2019); (ii) all individuals discharged from hospital in 2017-2019 where influenza was coded as the primary reason for hospitalisation and who were alive and under follow-up one week after discharge.

### Outcomes and covariates

The outcomes were (i) time to first hospitalisation or death (composite outcome); (ii) all-cause mortality; and (iii) time to first cause-specific hospitalisation or death. Hospitalisations were identified from linked SUS data. All-cause mortality was identified using date of death in the primary care record so that deaths before 2019 (in the influenza group) could be included (as linked ONS data were not available prior to 2019); concordance of death dates between primary care and linked ONS data has been shown to be high.^16^ Cause-specific outcomes were categorised based on ICD-10 codes into infections (ICD-10 codes beginning with “A”), cancers except non-melanoma skin cancer (C, except C44), endocrine/nutritional/metabolic (E), mental health and cognitive (F, G30 and X60-84), nervous system (G, except G30), circulatory (I), COVID-19/influenza/pneumonia/other lower respiratory tract infections (J09-22, U07.1/2), other respiratory (J23-99), digestive (K), musculoskeletal (M), genitourinary (N), and external causes (S-Y, except X60-84). For each of these, the outcome was time to the earliest of hospitalisation with the relevant outcome listed as primary diagnosis, or death with the relevant outcome listed as the underlying cause on the death certificate.^17^ The influenza control group was restricted to those discharged in 2019 for analyses of these cause-specific outcomes, because we did not have linked death registration data (and thus cause of death) for earlier years.

Other covariates considered in the analysis were factors that might be associated with both risk of severe COVID-19 and subsequent outcomes, namely age, sex, ethnicity, obesity, smoking status, index of multiple deprivation quintile (derived from the patient’s postcode at lower super output area level), and comorbidities considered potential risk factors for severe COVID-19 outcomes (see Table 1 and footnotes for full specification of covariate categories and comorbidities).

**Table 1:** Characteristics of patients hospitalised for COVID-19 and controls

|  |  | Hospitalised with<br>COVID-19 | Matched controls from<br>2019 general population | Hospitalised with influenza<br>in 2017-19 |
| --- | --- | --- | --- | --- |
| **N (%)** |  | 24673 (100.0) | 123362 (100.0) | 16058 (100.0) |
| **Age (yrs)** | 18-39 | 2035 (8.2) | 10175 (8.2) | 2024 (12.6) |
|  | 40-49 | 2756 (11.2) | 13780 (11.2) | 1462 (9.1) |
|  | 50-59 | 4679 (19.0) | 23395 (19.0) | 2126 (13.2) |
|  | 60-69 | 4602 (18.7) | 23010 (18.7) | 2653 (16.5) |
|  | 70-79 | 5034 (20.4) | 25170 (20.4) | 3492 (21.7) |
|  | 80+ | 5567 (22.6) | 27832 (22.6) | 4301 (26.8) |
|  | Median (IQR) | 66 (53-78) | 66 (53-78) | 69 (52-80) |
| **Sex** | Male | 13733 (55.7) | 68662 (55.7) | 7097 (44.2) |
|  | Female | 10940 (44.3) | 54700 (44.3) | 8961 (55.8) |
| **Body mass index (kg/m<sup>2</sup>)** | Not obese | 12710 (51.5) [54.8] | 82908 (67.2) [72.7] | 10065 (62.7) [67.4] |
| N (% of total) | 30-34.9 (Obese class I) | 5860 (23.8) [25.3] | 20985 (17.0) [18.4] | 2853 (17.8) [19.1] |
| [% among non-missing] | 35-39.9 (Obese class II) | 2819 (11.4) [12.2] | 7069 (5.7) [6.2] | 1271 (7.9) [8.5] |
|  | ≥40 (Obese class III) | 1789 (7.3) [7.7] | 3015 (2.4) [2.6] | 737 (4.6) [4.9] |
|  | Missing | 1495 (6.1) | 9385 (7.6) | 1132 (7.0) |
| **Smoking status** | Never | 10350 (41.9) [42.2] | 52145 (42.3) [43.0] | 5711 (35.6) [35.8] |
| N (% of total) | Former | 12498 (50.7) [51.0] | 52426 (42.5) [43.2] | 7346 (45.7) [46.1] |
| [% among non-missing] | Current | 1663 (6.7) [6.8] | 16699 (13.5) [13.8] | 2874 (17.9) [18.0] |
|  | Missing | 162 (0.7) | 2092 (1.7) | 127 (0.8) |
| **Ethnicity** | White | 19061 (77.3) [78.3] | 80923 (65.6) [87.7] | 14035 (87.4) [88.5] |
| N (% of total) | Mixed | 313 (1.3) [1.3] | 821 (0.7) [0.9] | 121 (0.8) [0.8] |
| [% among non-missing] | South Asian | 3457 (14.0) [14.2] | 6727 (5.5) [7.3] | 1242 (7.7) [7.8] |
|  | Black | 920 (3.7) [3.8] | 2225 (1.8) [2.4] | 251 (1.6) [1.6] |
|  | Other | 590 (2.4) [2.4] | 1572 (1.3) [1.7] | 211 (1.3) [1.3] |
|  | Missing | 332 (1.3) | 31094 (25.2) | 198 (1.2) |
| **Index of Multiple Deprivation** | 1 (least deprived) | 4622 (18.7) | 25428 (20.6) | 3282 (20.4) |
|  | 2 | 4743 (19.2) | 25259 (20.5) | 3251 (20.2) |
|  | 3 | 4678 (19.0) | 23503 (19.1) | 3272 (20.4) |
|  | 4 | 5183 (21.0) | 24222 (19.6) | 3133 (19.5) |
|  | 5 (most deprived) | 5447 (22.1) | 24950 (20.2) | 3120 (19.4) |
| **Care home resident** | Yes | 1197 (4.9) | 1650 (1.3) | 391 (2.4) |
| **Length of hospital stay** | Median (IQR) | 7 (3-13) | - | 4 (2-9) |
| **Any critical care** | Yes | 2659 (10.8) | - | 18 (0.1) |
| **Comorbidities** |  |  |  |  |
| Hypertension |  | 12132 (49.2) | 48565 (39.4) | 7550 (47.0) |
| Chronic respiratory disease |  | 3841 (15.6) | 9664 (7.8) | 3588 (22.3) |
| Asthma | With no oral steroid use | 3741 (15.2) | 14364 (11.6) | 2872 (17.9) |
|  | With oral steroid use | 1334 (5.4) | 2375 (1.9) | 1210 (7.5) |
| Chronic heart disease |  | 5540 (22.5) | 18285 (14.8) | 3934 (24.5) |
| Diabetes | With HbA1c<58 mmol/mol | 4727 (19.2) | 14855 (12.0) | 2443 (15.2) |
|  | With HbA1c≥58 mmol/mol | 3124 (12.7) | 5567 (4.5) | 1426 (8.9) |
|  | With no recent HbA1c measure | 402 (1.6) | 1133 (0.9) | 218 (1.4) |
| Cancer (non-haematological) | Diagnosed < 1 year ago | 401 (1.6) | 1044 (0.8) | 316 (2.0) |
|  | Diagnosed 1-4.9 years ago | 708 (2.9) | 2959 (2.4) | 539 (3.4) |
|  | Diagnosed ≥5 years ago | 1622 (6.6) | 7353 (6.0) | 1167 (7.3) |
| Haematological malignancy | Diagnosed < 1 year ago | 70 (0.3) | 123 (0.1) | 110 (0.7) |
|  | Diagnosed 1-4.9 years ago | 167 (0.7) | 362 (0.3) | 239 (1.5) |
|  | Diagnosed ≥5 years ago | 252 (1.0) | 694 (0.6) | 295 (1.8) |
| Reduced kidney function | Estimated GFR 30-60 | 4502 (18.2) | 17986 (14.6) | 3299 (20.5) |
|  | Estimated GFR 15-<30 | 481 (1.9) | 1313 (1.1) | 350 (2.2) |
|  | Estimated GFR <15 or dialysis | 443 (1.8) | 353 (0.3) | 342 (2.1) |
| Chronic liver disease |  | 414 (1.7) | 901 (0.7) | 222 (1.4) |
| Dementia |  | 1677 (6.8) | 4409 (3.6) | 1198 (7.5) |
| Stroke |  | 1835 (7.4) | 4275 (3.5) | 1057 (6.6) |
| Other neurological disease |  | 861 (3.5) | 1817 (1.5) | 574 (3.6) |
| Organ transplant |  | 173 (0.7) | 168 (0.1) | 189 (1.2) |
| Asplenia |  | 99 (0.4) | 242 (0.2) | 78 (0.5) |
| Rheum arthritis/ lupus /psoriasis |  | 2132 (8.6) | 7717 (6.3) | 1408 (8.8) |
| Other immunosuppressive disease |  | 76 (0.3) | 311 (0.3) | 108 (0.7) |

Note: Diabetes HbA1c category was determined according to the most recent glycated haemoglobin (HbA1c) recorded in the 15 months prior to the index date; other neurological disease was defined as motor neurone disease, myasthenia gravis, multiple sclerosis, Parkinson's disease, cerebral palsy, quadriplegia or hemiplegia, and progressive cerebellar disease; asplenia included splenectomy or a spleen dysfunction, including sickle cell disease; other immunosuppressive conditions was defined as permanent immunodeficiency ever diagnosed, or aplastic anaemia or temporary immunodeficiency recorded within the last year.

Information on all covariates was obtained by searching TPP SystmOne records for specific coded data, based on a subset of SNOMED-CT mapped to Read version 3 codes. Covariates were identified using data prior to the patient’s hospital admission date (for the COVID-19 and influenza groups) or the index date (for the matched control group, i.e. 1^st^ day of the matched calendar month in 2019). For the COVID-19 and influenza hospitalised groups, primary care data on ethnicity was supplemented with information from the hospitalisation record, to improve completeness. All codelists, along with detailed information on their compilation are available at https://codelists.opensafely.org for inspection and re-use by the wider research community.

### Statistical analysis

Follow-up began on the 8^th^ day after hospital discharge for the COVID-19 and influenza groups, and on the 1^st^ of the same calendar month in 2019, for the general population control group. Follow-up ended at the first occurrence of the analysis-specific outcome, or the earliest relevant censoring date for data availability/coverage for the outcome being analysed; the control groups were additionally censored after the maximum follow-up time of the COVID-19 group (315 days). For outcomes involving hospital admissions the administrative censoring date (for SUS data) was 30^th^ December 2020; for outcomes involving cause of death, the administrative censoring date (for ONS mortality data) was 11^th^ March 2021; for the all-cause death outcome which was ascertained in primary care data, patients were censored at date of deregistration if they had left the TPP general practice network. Cumulative incidence of the composite hospitalisation/death outcome and all-cause mortality were calculated using Kaplan-Meier methods. Hazard ratios comparing COVID-19 and controls were estimated using Cox regression models. Separate models were fitted for the comparisons with matched 2019 general population controls (models stratified by matched set), and with influenza controls (models adjusted for age, sex, STP and calendar month). The additional covariates noted above were then added to the models. People with missing data on ethnicity, body mass index or smoking were excluded from models that used these variables (“complete case analysis”, which is valid under the assumption that missingness is conditionally independent of the outcome);^18^ imputation was not used because these variables were thought to be missing not at random in primary care (e.g. smokers more likely to have smoking status recorded). Cumulative incidence of cause-specific hospitalisation/death outcomes were calculated with deaths from other causes treated as a competing risk. Hazard ratios for these outcomes were then estimated from a Cox model targeting the cause-specific hazard, with deaths from competing risks censored.

The study was approved by the Health Research Authority (REC reference 20/LO/0651) and by the LSHTM Ethics Board (ref 21863).

## Results

24,673 individuals discharged after a COVID-19 hospitalisation were included, alongside 123,362 matched controls from the 2019 general population, and 16,058 individuals discharged after influenza hospitalisation in 2017-19 (Appendix Figure A1-A2).

At entry, the COVID-19 group had similar age and sex distribution to the general population groups due to matching, but had younger median age and were more likely to be male than the influenza group (Table 1). Body mass index, smoking and ethnicity data were 93-99% complete in all groups, except that ethnicity was 25% missing in the matched control group (no hospital-based ethnicity records were available for this group). The COVID-19 group were more likely to be obese, non-White and less likely to be current smokers than both comparison groups. Pre-existing comorbidities were more common in both COVID-19 and influenza-discharged patients than general population controls. COVID-19 patients had longer median duration of hospital stay and were more likely to have received critical care during their admission than influenza patients.

Numbers of outcome events are shown in Appendix Table A1. Cumulative incidence of subsequent hospital admission or death after study entry in the COVID-19 group was higher than in general population controls, but similar to that in the influenza group (cumulative incidence at 6 months = 34.8, 15.2 and 37.8% in the three groups respectively; fully adjusted HR = 2.23, 2.14-2.31 for COVID-19 vs general population; 0.94, 0.91-0.98 for COVID-19 vs influenza, Figures 1a, 2). Cumulative all-cause mortality was higher in the COVID-19 group than both the general population and influenza groups (7.5, 1.4 and 4.9% at 6 months in the three groups respectively; fully adjusted HR = 4.97, 4.58-5.40 for COVID-19 vs general population; 1.73, 1.60-1.87 for COVID vs influenza, Figures 1b, 2). To further explore this, causes of death were examined (appendix Table A2). A substantial proportion of deaths in the COVID-19 group had COVID-19 listed as the underlying cause (500/2022, 24.7%), while in the influenza group ≤5 deaths were coded with influenza as the underlying cause.

**Figure 1:**
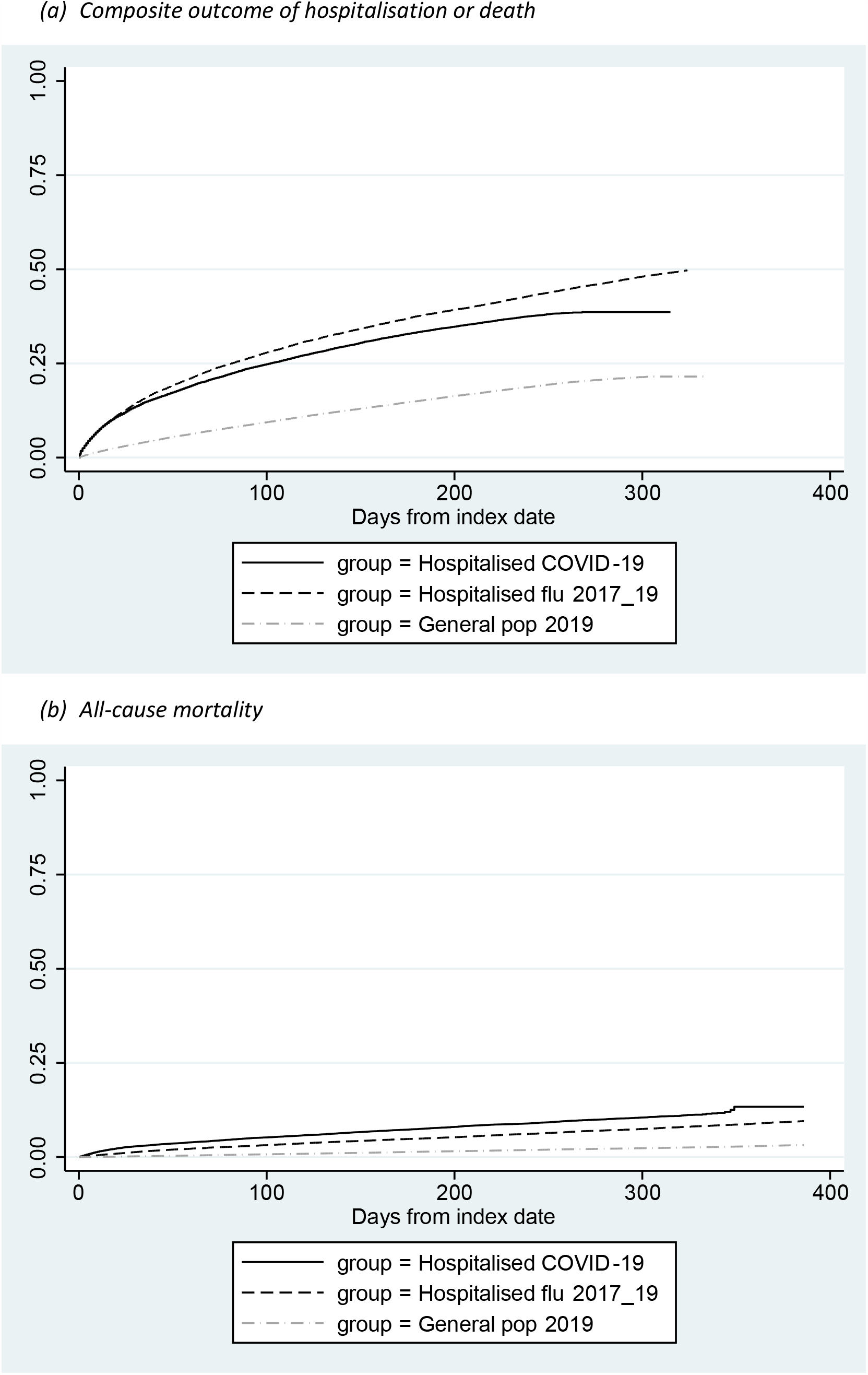
Cumulative incidence of admission or death (composite outcome), and all-cause mortality in patients discharged from COVID-19 hospital admissions, influenza hospital admissions, and in matched general population controls

**Figure 2:**
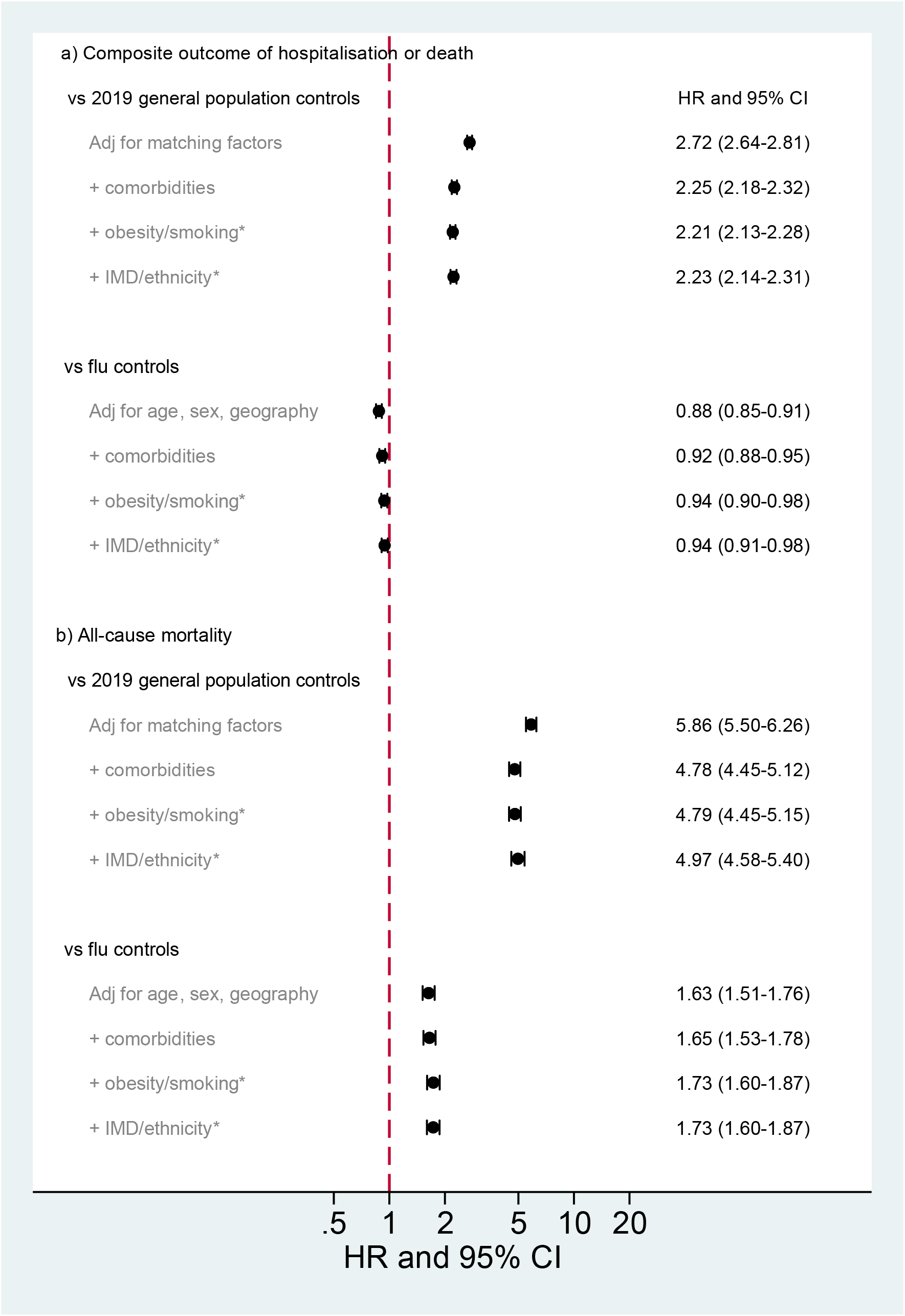
Hazard ratios comparing exposed (prior COVID-19 hospitalisation) and controls for risk of subsequent hospital admission or death (composite outcome) and all-cause mortality *among those with complete data available (see Table 1)

Cumulative incidences of cause-specific hospital admissions or deaths are shown in Figure 3. After adjustment for matching factors and other covariates, risks of all cause-specific outcomes were substantially higher in COVID-19 groups than general population controls (Figure 4). Compared with influenza patients, people in the COVID-19 group had similar or lower risk of admission or death from most causes, but higher risks of admission/death from COVID-19/influenza/lower respiratory tract infection (LRTI, adjusted HR 1.37, 1.22-1.54); in the post-COVID-19 group these outcomes were dominated by codes for COVID-19 itself (515/1122 [46%] of hospitalisations and 342/368 [93%] of deaths) and pneumonia (461/1122 [41%] of hospitalisations). The COVID-19 group also had higher risks than the influenza group for mental health or cognitive outcomes (adjusted HR 1.36, 1.01-1.83). This was further explored in a post-hoc analysis of specific outcomes within the mental health and cognitive category (Table 2). Raised risks in the COVID-19 group appeared to be driven by dementia hospitalisations/deaths (age/sex-adjusted HR 2.32, 1.48-3.64), particularly among those with pre-existing dementia at baseline (HR 2.47, 1.37-4.44) and/or resident in care homes (HR 2.53, 0.99-6.41). Of note, 129/161 dementia outcome events (80.1%) were deaths (rather than hospitalisations). Higher rates of hospitalisations/deaths due to mood disorders, and neurotic/stress-related/somatoform disorders were also observed in COVID-19 patients, but confidence intervals were too wide to be conclusive.

**Figure 3:**
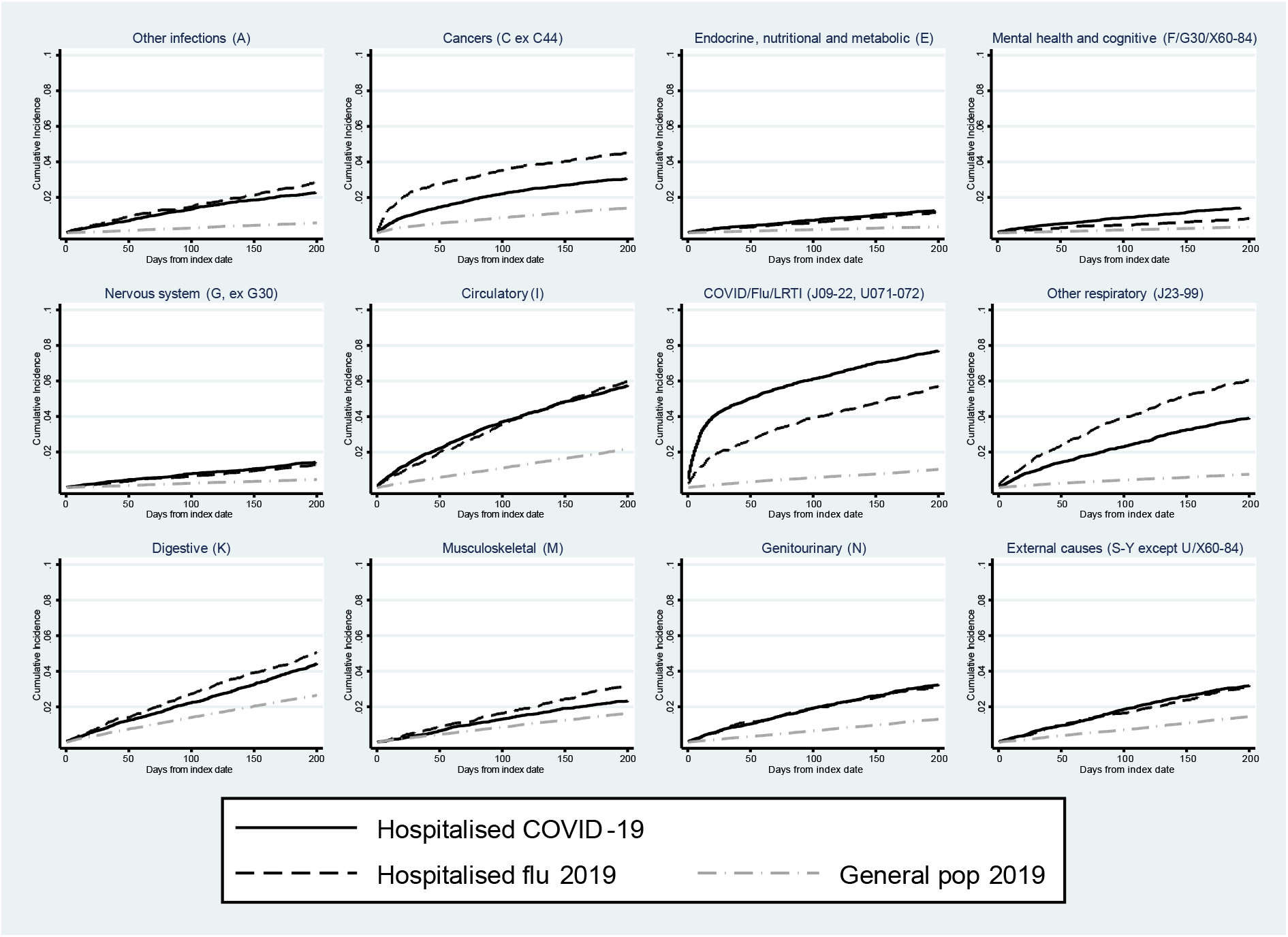
Cumulative incidence of cause-specific admission/death in patients discharged from COVID-19 hospital admissions, influenza hospital admissions, and in matched general population controls Note: For each sub-panel in (c), the outcome was defined as the first hospitalisation or death record with an ICD-10 code in the given category listed as the primary reason for hospitalisation/underlying cause of death. Deaths from other causes were treated as competing risks. In the influenza group, only patients entering the study in 2019 were included in analysis of cause-specific outcomes, as linked cause of death data were only available from 2019 onwards.

**Figure 4:**
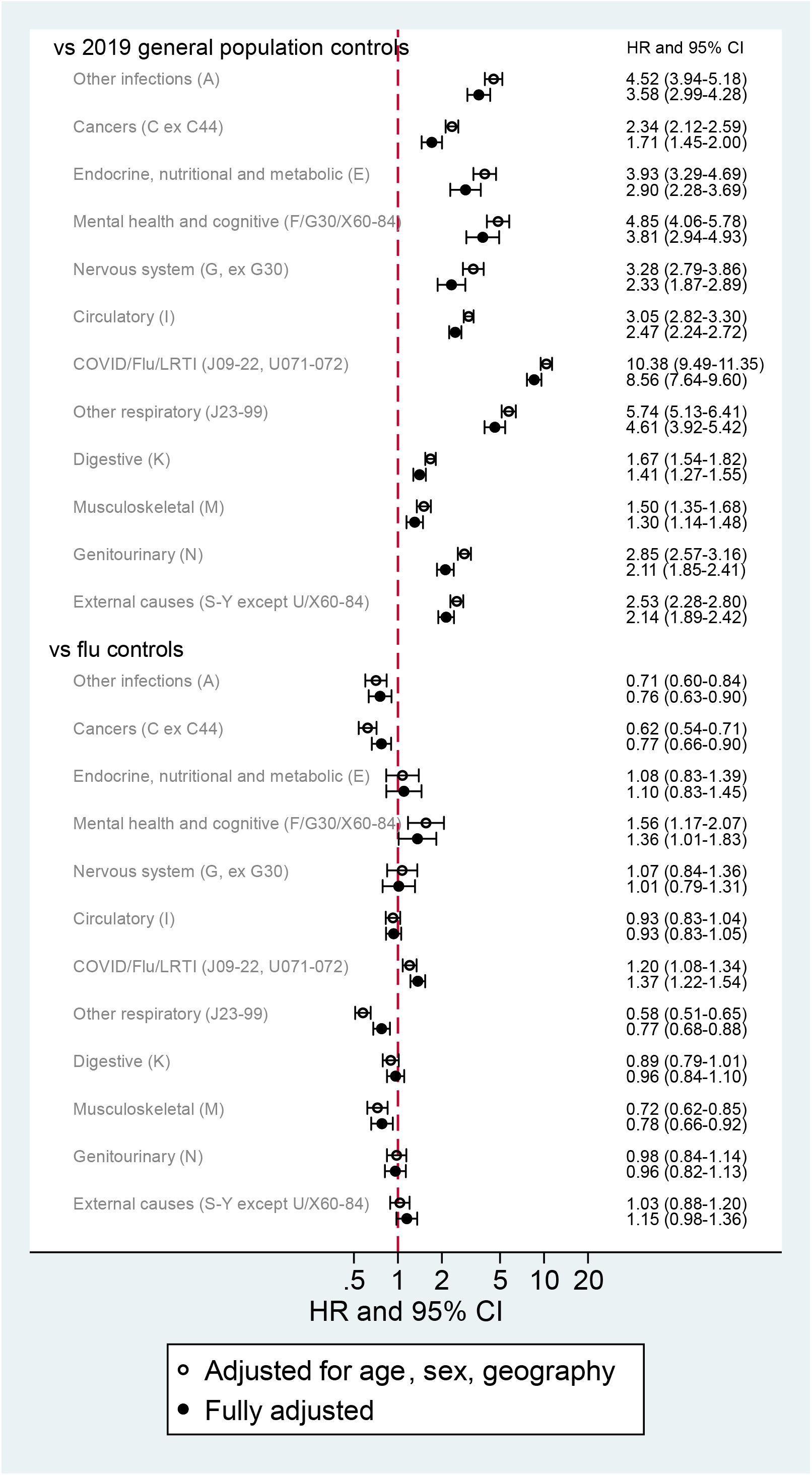
Hazard ratios comparing exposed (prior COVID-19 hospitalisation) and controls for cause-specific hospital admission/deaths, adjusted for age, sex and geography *In the influenza group, only patients entering the study in 2019 were included in analysis of cause-specific outcomes, as linked cause of death data were only available from 2019 onwards.

**Table 2:** Post-hoc analysis of specific hospitalisation/mortality outcomes within the mental health and cognitive category

|  | [events] rate per 1000 person-years (95% CI) |  | Age- and sex- adjusted<br>hazard ratio<br>for COVID-19 vs<br>influenza groups<br>(95% CI) |
| --- | --- | --- | --- |
|  | COVID-19 group | Influenza group |  |
| **Dementia (F00-F03, G30)** | [134] 14.74 (12.45-17.46) | [27] 5.20 (3.57-7.58) | 2.32 (1.48-3.64) |
| *(among those with baseline dementia)* | [102] 168.07 (138.43-204.07) | [14] 59.72 (35.37-100.84) | 2.47 (1.37-4.44) |
| *(among those with no baseline dementia)* | [32] 3.77 (2.67-5.33) | [13] 2.62 (1.52-4.52) | 1.23 (0.63-2.42) |
| *(among those resident in a care home)* | [56] 127.78 (98.34-166.04) | [<=5] 55.14 (22.95-132.47) | 2.53 (0.99-6.41) |
| *(among those not resident in a care home)* | [78] 9.02 (7.22-11.26) | [22] 4.31 (2.84-6.55) | 1.80 (1.08-2.98) |
| **Delirium (F05)** | [77] 8.47 (6.78-10.59) | [28] 5.39 (3.72-7.81) | 1.10 (0.70-1.72) |
| **Schizophrenia/schizotypal/**<br>**delusional disorders (F20-29)** | [<=5] 0.22 (0.06-0.88) | [<=5] 0.77 (0.29-2.05) | 0.20 (0.03-1.12) |
| **Mood disorders (F30-39)** | [19] 2.09 (1.33-3.28) | [<=5] 0.77 (0.29-2.05) | 1.61 (0.54-4.79) |
| **Neurotic/stress-related/**<br>**somatoform disorders (F40-48)** | [13] 1.43 (0.83-2.46) | [<=5] 0.58 (0.19-1.79) | 2.59 (0.71-9.52) |
| **All except dementia (F05-F99)** | [114] 12.54 (10.44-15.07) | [41] 7.90 (5.82-10.73) | 1.12 (0.77-1.62) |

## Discussion

### Key findings

Patients discharged from a COVID-19 hospitalisation had more than double the risk of hospitalisation or death, and a 5-fold higher risk of all-cause mortality than controls from the general population, after adjusting for baseline personal and clinical characteristics. Risks were higher for all categories of disease-specific hospital admissions/deaths after a COVID-19 hospitalisation than in general population controls. Most outcomes were similar between people discharged from a COVID-19 hospitalisation, and people discharged from an influenza hospitalisation in 2017-19, but the COVID-19 group had higher subsequent all-cause mortality; higher rates of respiratory infection admissions and deaths (predominantly COVID-19), and more adverse mental health and cognitive outcomes (particularly deaths attributed to dementia among people with pre-existing dementia) compared with the influenza group.

### Findings in context of literature and explanations

A recent study of VA data on US veterans examined a wide range of diagnostic and other outcomes in 30-day COVID-19 survivors, compared with the general VA population.^13^ Among veterans where COVID-19 had led to a hospitalisation, hazard ratios of every category of outcome were raised. This concurs with findings from our study, despite different characteristics of the VA population. In the UK, an earlier study found an 8-fold higher risk of death in post-acute COVID-19 patients compared with general population controls, and raised risks of respiratory disease, diabetes and cardiovascular disease.^15^ Interestingly, recent data from Denmark suggest limited post-acute complications following non-hospitalised COVID-19;^19^ this is in contrast to a recent study using US health insurance data which found raised risks of a range of outcomes among a relatively young cohort with mostly (92%) non-hospitalised COVID-19 disease, compared with both the general population and people with a record of other viral lower respiratory tract infections.^14^

Our data showed that COVID-19 hospitalised patients were more likely to have baseline comorbidities than general population controls, reflecting known associations between comorbidities and risks of severe COVID-19 outcomes.^6^ Differences in outcomes between hospitalised patients and general population controls might therefore reflect baseline differences not fully captured in our adjustment models, and might also reflect a generic adverse effect of hospitalisation.^20^ This is supported by the more similar risks we observed when COVID-19 survivors were compared with people who had experienced influenza hospitalisation. However, all-cause mortality was substantially higher after COVID-19 compared with influenza. A quarter of deaths after a COVID-19 hospitalisation had COVID-19 listed as the underlying cause, but it is not clear from our data whether patients experienced specific complications after hospital discharge that were then attributed to COVID-19.

Our analysis of cause-specific outcomes also suggested a disproportionate rate of dementia deaths post-COVID-19, particularly among those with pre-existing dementia. Cognitive decline after hospitalisation and critical illness have been previously described;^21,22^ acute COVID-19 and associated hospital admission, social isolation, and medications may have accelerated progression of patients’ dementia; it is unclear whether post-discharge care was adequate for this vulnerable group. However, it is possible that deaths where the underlying cause was recorded as dementia may have been due to progression of underlying health problems following an acute illness as well as difficulty in managing these due to dementia. Due to small numbers, we could not confirm whether higher rates of mood disorders and neurotic/stress-related/somatoform disorders after COVID-19 compared with influenza were due to chance, but a number of previous studies outside the pandemic context have found that critical illness is associated with raised risks of depression, anxiety and post-traumatic stress.^23-25^ It will be important to continue to monitor these outcomes as more follow-up accumulates.

### Strengths and Limitations

We identified COVID-19 hospitalisations from a base population covering around 40% of the population of England, giving us high statistical power. We examined a broad range of hospitalisation and mortality outcomes, and were able to describe and adjust for a wide range of personal and clinical characteristics using rich primary care data.

However, our study has some limitations. We relied on ICD-10 codes entered as the primary reason for hospitalisation or underlying cause of death to define our cause-specific outcomes, but these fields may not have been used consistently.^26^ In particular, there might have been a tendency for clinicians aware of a recent COVID-19 hospitalisation to code COVID-19 for a range of clinical complications, masking more specific sequelae. Outcomes were classified in broad categories to obtain an overview of post-COVID-19 disease patterns; more granular disease categories would be of future interest but will require more follow-up to maintain statistical power. Our main comparisons may have been affected by time-related factors. We compared post-COVID patients in 2020 with controls from 2019 and earlier; consultations for non-COVID-19 conditions in 2020 are known to have been subdued in the general population,^27^ perhaps due to lockdown, or public reluctance to seek care, potentially affecting comparison with earlier years. On the other hand, patients with recent a COVID-19 hospitalisation may assume immunity from reinfection and be less reticent in seeking care than the general population. The comparison with influenza may also have been affected by seasonality, since the first wave of COVID-19 in England happened outside the typical influenza season. Lack of overlap in the data meant that we could not incorporate seasonal adjustment into our statistical models for this comparison; any confounding by seasonality is likely to have led to underestimation of hazard ratios comparing COVID-19 and influenza patients, since cases of the former were under-represented in the winter months (which typically confer higher health risks). We did not have detailed data on disease severity, though descriptive data showed that COVID-19 patients tended to have longer hospital stays and more critical care than those hospitalised for influenza. Finally, data were unavailable on new/emerging COVID-19 variants during the study period.

## Conclusions

Patients surviving a COVID-19 hospitalisation were at substantially higher risk than the general population for a range of subsequent adverse outcomes over a period of up to 10 months’ follow-up included in this study. Risks for most outcomes were broadly comparable to those experienced by influenza hospitalisation survivors prior to the pandemic, but in the period following hospital discharge COVID-19 patients had higher risks of all-cause mortality, readmission or death attributed to their initial infection, and adverse mental health and cognitive outcomes; in particular, among people with pre-existing dementia, we observed an excess of deaths where dementia was recorded as the underlying cause. These findings suggest a need for services to support and closely monitor people following discharge from hospital with COVID-19.

## Data Availability

All data were linked, stored and analysed securely within the OpenSAFELY platform https://opensafely.org/. Data include pseudonymized data such as coded diagnoses, medications and physiological parameters. No free text data are included. All code is shared openly for review and re-use under MIT open license (https://github.com/opensafely/post-admission-admissions-research). Detailed pseudonymised patient data is potentially re-identifiable and therefore not shared. We rapidly delivered the OpenSAFELY data analysis platform without prior funding to deliver timely analyses on urgent research questions in the context of the global Covid-19 health emergency: now that the platform is established we are developing a formal process for external users to request access in collaboration with NHS England; updates on this process are available on OpenSAFELY.org.

## Contributors

KB, CTR, WJH led the study design. GH led on platform support. KB did the analysis and wrote the first draft. BG conceived the OpenSAFELY platform and the approach. BG and LS lead the project overall and are guarantors. SB led on software development. AM led on information governance. Other contributions were – data curation - CB, JP, JC, SH, SB, DE, PI and CEM; disease category conceptualisation and codelists - KB CTR BM CB JC CEM AJW HIM IJD HJC JP; statistical analysis code: KB, EW; ethical approvals - HJC EW LS BG; software - SB, DE, PI, AJW, WJH, CEM, CB, FH, JC; writing (reviewing and editing) - KB, CTR, CWG, LT, BM, AS, AM, RME, CEM, IJD, SJWE, LS, BG. All authors were involved in design and conceptual development and reviewed and approved the final manuscript.

## Information Governance and Ethics

NHS England is the data controller; TPP is the data processor; and the key researchers on OpenSAFELY are acting on behalf of NHS England. OpenSAFELY is hosted within the TPP environment which is accredited to the ISO 27001 information security standard and is NHS IG Toolkit compliant;^28,29^ patient data are pseudonymised for analysis and linkage using industry standard cryptographic hashing techniques; all pseudonymised datasets transmitted for linkage onto OpenSAFELY are encrypted; access to the platform is via a virtual private network (VPN) connection, restricted to a small group of researchers who hold contracts with NHS England and only access the platform to initiate database queries and statistical models. All database activity is logged. No patient-level data leave the platform; only aggregate statistical outputs leave the platform environment following best practice for anonymisation of results such as statistical disclosure control for low cell counts.^30^ The OpenSAFELY platform adheres to the data protection principles of the UK Data Protection Act 2018 and the EU General Data Protection Regulation (GDPR) 2016. In March 2020, the Secretary of State for Health and Social Care used powers under the UK Health Service (Control of Patient Information) Regulations 2002 (COPI) to require organisations to process confidential patient information for the purposes of protecting public health, providing healthcare services to the public and monitoring and managing the COVID-19 outbreak and incidents of exposure.^31^ Taken together, these provide the legal bases to link patient datasets on the OpenSAFELY platform. The study was approved by the Health Research Authority (REC reference 20/LO/0651) and by the LSHTM Ethics Board (ref 21863).

## Open access statement

This research was funded in whole or in part by the Wellcome Trust. For the purpose of Open Access, the author has applied a CC BY public copyright licence to any Author Accepted Manuscript (AAM) version arising from this submission.

## Patient and Public Involvement

Patients were not formally involved in developing this specific study design that was developed rapidly in the context of a global health emergency. We have developed a publicly available website https://opensafely.org/ through which we invite any patient or member of the public to contact us regarding this study or the broader OpenSAFELY project.

## Appendix

**Figure A1:**
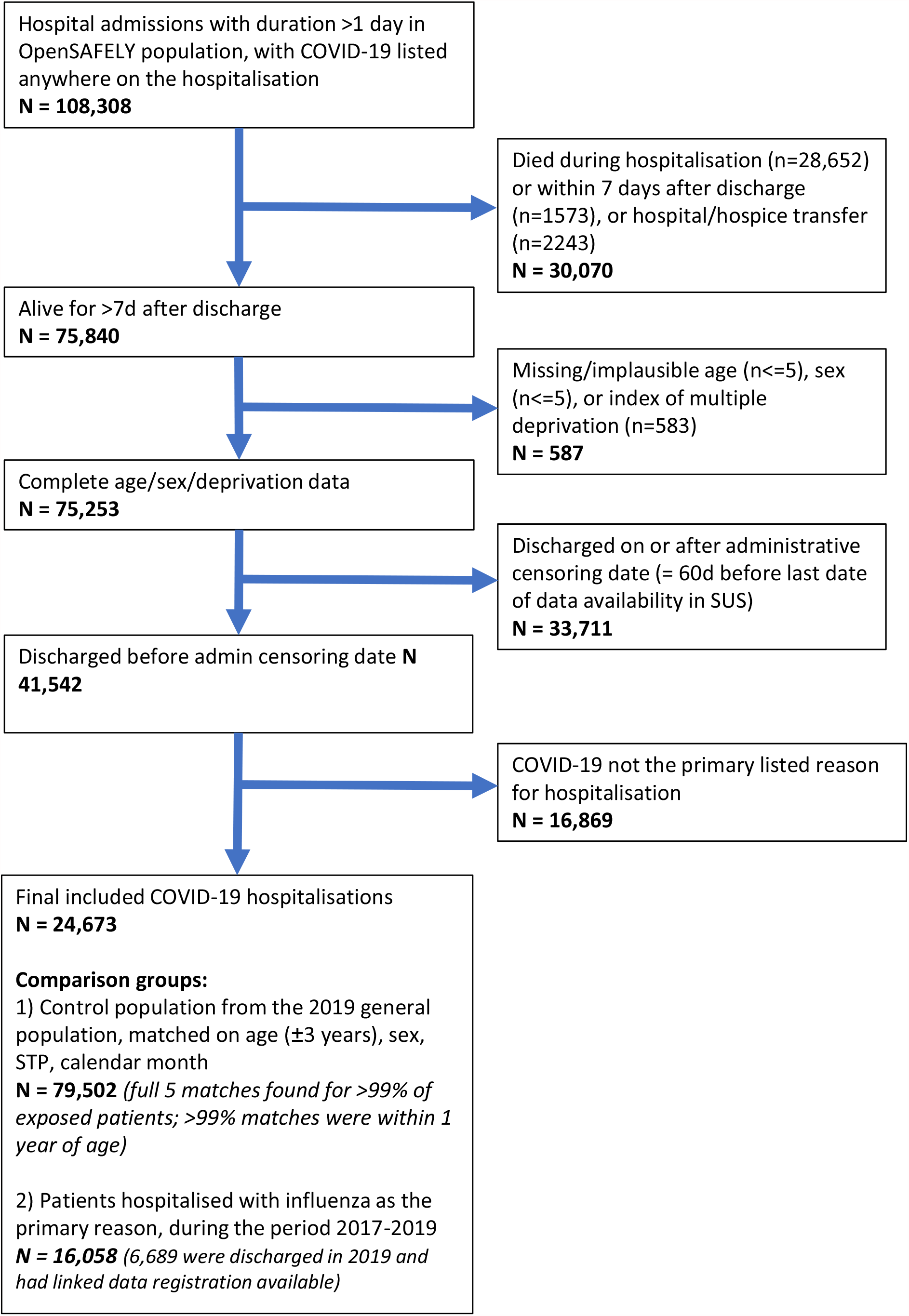
Study flow chart

**Figure A2:**
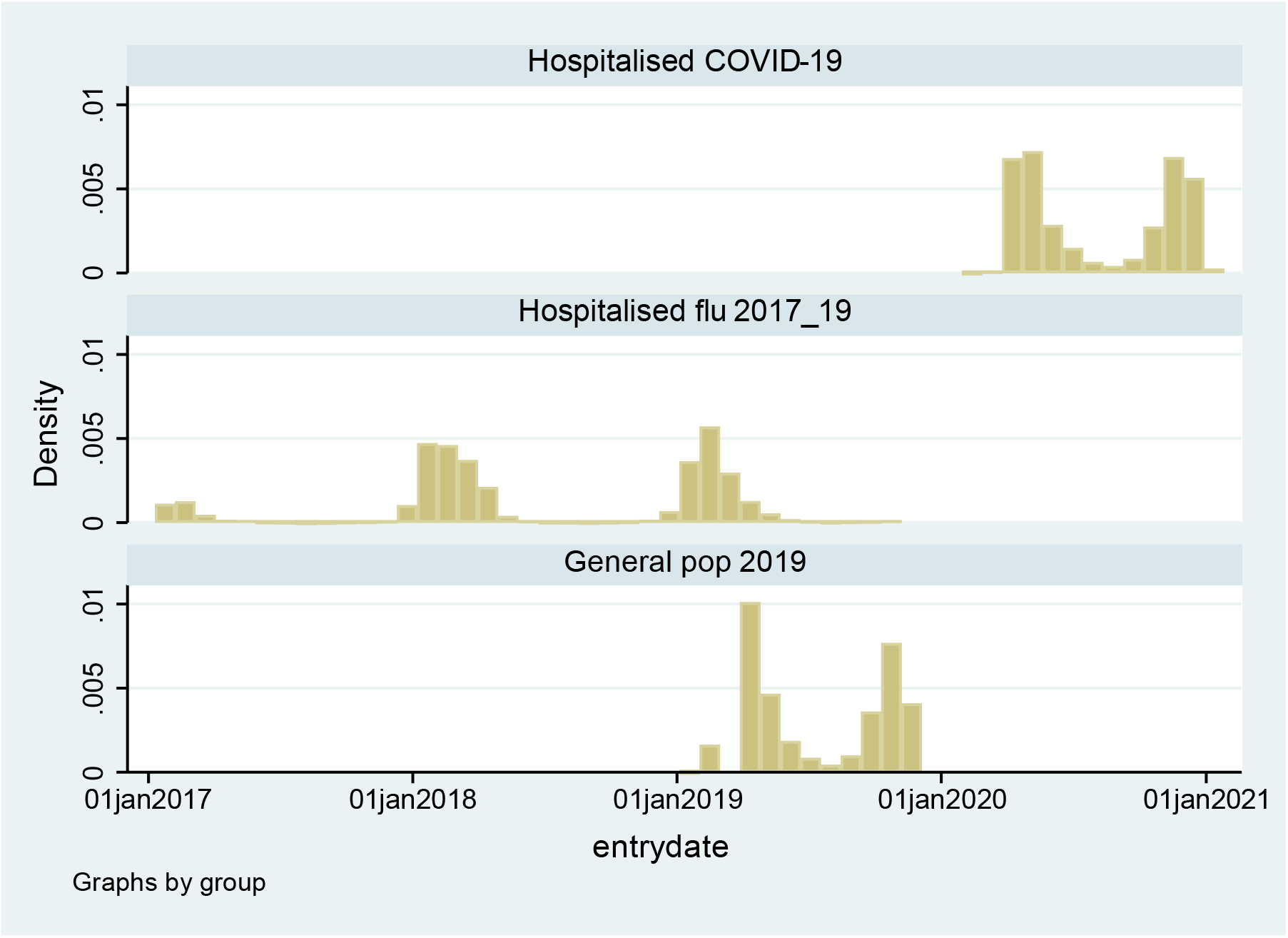
Distribution of entry dates for those in the COVID-19 hospitalised group, and the influenza-hospitalised and matched general population comparison groups

**Table A1:** Distribution of first outcomes (hospital admission or death) among included individuals

|  | Hospitalised with<br>COVID-19 | Hospitalised with<br>influenza in<br>2017-19* | Matched controls<br>from 2019 general<br>population |
| --- | --- | --- | --- |
| N | 24673 | 16058 (6689 in<br>2019*) | 123362 |
| Any hospitalisation or death (composite outcome) | 6499 (26.3) | 7788 (48.5) | 16681 (13.5) |
| Deaths | 2022 (8.2) | 1513 (9.4) | 3904 (3.2) |
| **Cause-specific admissions/deaths**<br>**(total [deaths])** |  |  |  |
| Other infections (ICD-10 codes beginning A) | 391 [<=5] (1.6) | 255 [<=5] (3.8) | 569 [6] (0.5) |
| Cancers (C, ex C44) | 566 [174] (2.3) | 355 [60] (5.3) | 1478 [242] (1.2) |
| Endocrine, nutritional and metabolic (E) | 218 [29] (0.9) | 98 [13] (1.5) | 361 [28] (0.3) |
| Mental health and cognitive (F/G30/X60-84) | 248 [111] (1.0) | 68 [26] (1.0) | 356 [149] (0.3) |
| Nervous system (G, ex G30) | 240 [84] (1.0) | 116 [21] (1.7) | 467 [98] (0.4) |
| Circulatory (I) | 1039 [180] (4.2) | 511 [56] (7.6) | 2256 [300] (1.8) |
| COVID-19/Influenza/LRTI (J09-22, U07.1/2) | 1490+ [368+] (6.0) | 497 [18] (7.4) | 1169 [55] (0.9) |
| Other respiratory (J23-99) | 670 [98] (2.7) | 526 [40] (7.9) | 812 [100] (0.7) |
| Digestive (K) | 751 [36] (3.0) | 433 [11] (6.5) | 2728 [26] (2.2) |
| Musculoskeletal (M) | 408 [9] (1.7) | 292 [7] (4.4) | 1669 [14] (1.4) |
| Genitourinary (N) | 573 [18] (2.3) | 287 [<=5] (4.3) | 1292 [28] (1.0) |
| External (S-Y, ex X60-84) | 566 [<=5] (2.3) | 301 [<=5] (4.5) | 1476 [16] (1.2) |

Note: counts for cause specific deaths in square brackets are deaths which were not preceded by a previous hospitalisation in the same outcome category; these are the deaths that counted as outcome events in the cause-specific analysis, and not the total number of deaths from the given cause. \*In the influenza group, only patients entering the study in 2019 were included in analysis of cause-specific outcomes, as linked cause of death data were only available from 2019 onwards. \*In the COVID-19 group, 515/1122 (46%) hospitalisations in the COVID-19/Influenza/LRTI outcome category had COVID-19 codes (ICD-10 U07.1/2) and a further 461/1122 (41%) had pneumonia codes (J18) as the primary diagnosis; 342/368 deaths (93%) had COVID-19 as the underlying cause.

**Table A2:**
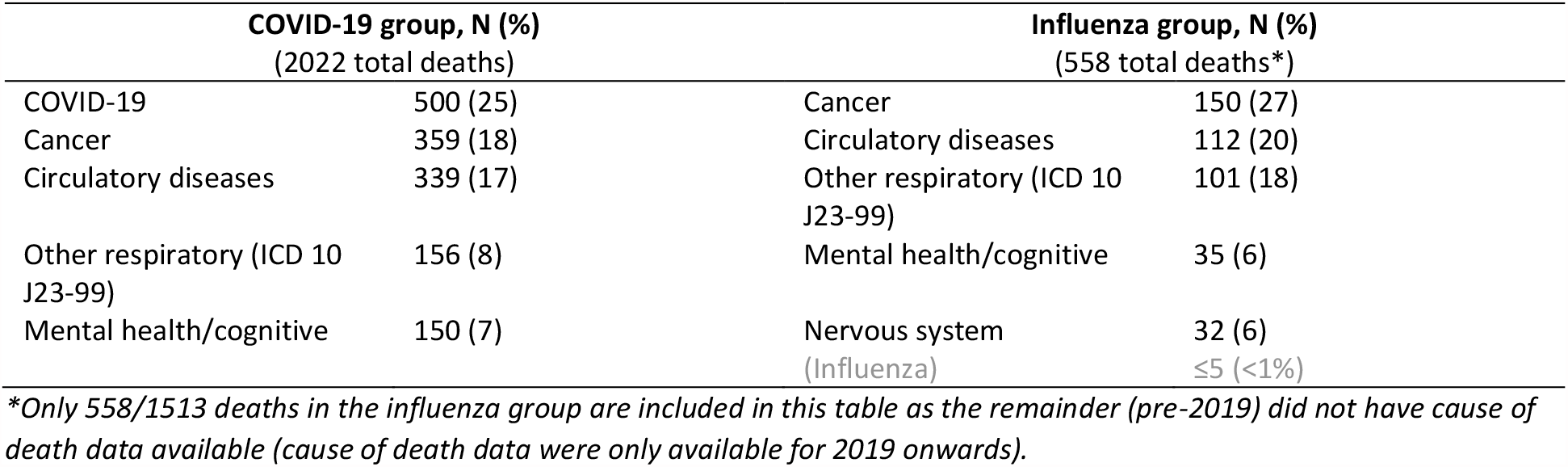
Leading causes of death in COVID-19 and influenza groups

## References

1. World Health Organization. WHO Coronavirus (COVID-19) Dashboard. 2021. https://covid19.who.int (accessed 12th May 2021).

2. Oran DP, Topol EJ. Prevalence of Asymptomatic SARS-CoV-2 Infection. Ann Intern Med 2020; 173(5): 362–7. doi: 10.7326/M20-3012.

3. Prieto-Alhambra D, Balló E, Coma E, Mora N, Aragón M, Prats-Uribe A, Fina F, Benítez M, Guiriguet C, Fàbregas M, Medina-Peralta M, Duarte-Salles T. Filling the gaps in the characterization of the clinical management of COVID-19: 30-day hospital admission and fatality rates in a cohort of 118 150 cases diagnosed in outpatient settings in Spain. Int J Epidemiol 2020; 49(6): 1930–9. doi: 10.1093/ije/dyaa190.

4. Clift AK, Coupland CAC, Keogh RH, Diaz-Ordaz K, Williamson E, Harrison EM, Hayward A, Hemingway H, Horby P, Mehta N, Benger J, Khunti K, Spiegelhalter D, Sheikh A, Valabhji J, Lyons RA, Robson J, Semple MG, Kee F, Johnson P, Jebb S, Williams T, Hippisley-Cox J. Living risk prediction algorithm (QCOVID) for risk of hospital admission and mortality from coronavirus 19 in adults: national derivation and validation cohort study. BMJ 2020; 371: m3731. doi: 10.1136/bmj.m3731.

5. Mathur R, Rentsch CT, Morton CE, Hulme WJ, Schultze A, MacKenna B, Eggo RM, Bhaskaran K, Wong AYS, Williamson EJ, Forbes H, Wing K, McDonald HI, Bates C, Bacon S, Walker AJ, Evans D, Inglesby P, Mehrkar A, Curtis HJ, DeVito NJ, Croker R, Drysdale H, Cockburn J, Parry J, Hester F, Harper S, Douglas IJ, Tomlinson L, Evans SJW, Grieve R, Harrison D, Rowan K, Khunti K, Chaturvedi N, Smeeth L, Goldacre B. Ethnic differences in SARS-CoV-2 infection and COVID-19-related hospitalisation, intensive care unit admission, and death in 17 million adults in England: an observational cohort study using the OpenSAFELY platform. The Lancet 2021; 397(10286): 1711–24. doi: 10.1016/S0140-6736(21)00634-6.

6. Williamson EJ, Walker AJ, Bhaskaran K, Bacon S, Bates C, Morton CE, Curtis HJ, Mehrkar A, Evans D, Inglesby P. OpenSAFELY: factors associated with COVID-19 death in 17 million patients. Nature 2020: 1–11. doi: 10.1038/s41586-020-2521-4.

7. Zhou F, Yu T, Du R, Fan G, Liu Y, Liu Z, Xiang J, Wang Y, Song B, Gu X, Guan L, Wei Y, Li H, Wu X, Xu J, Tu S, Zhang Y, Chen H, Cao B. Clinical course and risk factors for mortality of adult inpatients with COVID-19 in Wuhan, China: a retrospective cohort study. Lancet 2020; 395(10229): 1054–62. doi: 10.1016/S0140-6736(20)30566-3.

8. Navaratnam AV, Gray WK, Day J, Wendon J, Briggs TWR. Patient factors and temporal trends associated with COVID-19 in-hospital mortality in England: an observational study using administrative data. The Lancet Respiratory Medicine 2021; 9(4): 397–406. doi: 10.1016/S2213-2600(20)30579-8.

9. Siemieniuk RA, Bartoszko JJ, Ge L, Zeraatkar D, Izcovich A, Kum E, Pardo-Hernandez H, Qasim A, Martinez JPD, Rochwerg B, Lamontagne F, Han MA, Liu Q, Agarwal A, Agoritsas T, Chu DK, Couban R, Cusano E, Darzi A, Devji T, Fang B, Fang C, Flottorp SA, Foroutan F, Ghadimi M, Heels-Ansdell D, Honarmand K, Hou L, Hou X, Ibrahim Q, Khamis A, Lam B, Loeb M, Marcucci M, McLeod SL, Motaghi S, Murthy S, Mustafa RA, Neary JD, Rada G, Riaz IB, Sadeghirad B, Sekercioglu N, Sheng L, Sreekanta A, Switzer C, Tendal B, Thabane L, Tomlinson G, Turner T, Vandvik PO, Vernooij RW, Viteri-García A, Wang Y, Yao L, Ye Z, Guyatt GH, Brignardello-Petersen R. Drug treatments for covid-19: living systematic review and network meta-analysis. BMJ 2020; 370: m2980. doi: 10.1136/bmj.m2980.

10. Nalbandian A, Sehgal K, Gupta A, Madhavan MV, McGroder C, Stevens JS, Cook JR, Nordvig AS, Shalev D, Sehrawat TS, Ahluwalia N, Bikdeli B, Dietz D, Der-Nigoghossian C, Liyanage-Don N, Rosner GF, Bernstein EJ, Mohan S, Beckley AA, Seres DS, Choueiri TK, Uriel N, Ausiello JC, Accili D, Freedberg DE, Baldwin M, Schwartz A, Brodie D, Garcia CK, Elkind MSV, Connors JM, Bilezikian JP, Landry DW, Wan EY. Post-acute COVID-19 syndrome. Nat Med 2021; 27(4): 601–15. doi: 10.1038/s41591-021-01283-z.

11. Mazza MG, De Lorenzo R, Conte C, Poletti S, Vai B, Bollettini I, Melloni EMT, Furlan R, Ciceri F, Rovere-Querini P, Benedetti F. Anxiety and depression in COVID-19 survivors: Role of inflammatory and clinical predictors. Brain Behav Immun 2020; 89: 594–600. doi: https://doi.org/10.1016/j.bbi.2020.07.037.

12. Xiong Q, Xu M, Li J, Liu Y, Zhang J, Xu Y, Dong W. Clinical sequelae of COVID-19 survivors in Wuhan, China: a single-centre longitudinal study. Clin Microbiol Infect 2021; 27(1): 89–95. doi: https://doi.org/10.1016/j.cmi.2020.09.023.

13. Al-Aly Z, Xie Y, Bowe B. High-dimensional characterization of post-acute sequalae of COVID-19. Nature 2021. doi: 10.1038/s41586-021-03553-9.

14. Daugherty SE, Guo Y, Heath K, Dasmariñas MC, Jubilo KG, Samranvedhya J, Lipsitch M, Cohen K. Risk of clinical sequelae after the acute phase of SARS-CoV-2 infection: retrospective cohort study. BMJ 2021; 373: 1098. doi: 10.1136/bmj.n1098.

15. Ayoubkhani D, Khunti K, Nafilyan V, Maddox T, Humberstone B, Diamond I, Banerjee A. Post-covid syndrome in individuals admitted to hospital with covid-19: retrospective cohort study. BMJ 2021; 372: n693. doi: 10.1136/bmj.n693.

16. Gallagher AM, Dedman D, Padmanabhan S, Leufkens HGM, de Vries F. The accuracy of date of death recording in the Clinical Practice Research Datalink GOLD database in England compared with the Office for National Statistics death registrations. Pharmacoepidemiol Drug Saf 2019; 28(5): 563–9. doi: 10.1002/pds.4747.

17. Office of National Statistics’ Death Certification Advisory group. Guidance for doctors completing medical certificates of cause of death in England and Wales: Office of National Statistics, 2020.

18. White IR, Carlin JB. Bias and efficiency of multiple imputation compared with complete-case analysis for missing covariate values. Stat Med 2010; 29(28): 2920–31. doi: 10.1002/sim.3944.

19. Lund LC, Hallas J, Nielsen H, Koch A, Mogensen SH, Brun NC, Christiansen CF, Thomsen RW, Pottegård A. Post-acute effects of SARS-CoV-2 infection in individuals not requiring hospital admission: a Danish population-based cohort study. The Lancet Infectious Diseases. doi: 10.1016/S1473-3099(21)00211-5.

20. Krumholz HM. Post-hospital syndrome--an acquired, transient condition of generalized risk. The New England journal of medicine 2013; 368(2): 100–2. doi: 10.1056/NEJMp1212324.

21. Pandharipande PP, Girard TD, Jackson JC, Morandi A, Thompson JL, Pun BT, Brummel NE, Hughes CG, Vasilevskis EE, Shintani AK, Moons KG, Geevarghese SK, Canonico A, Hopkins RO, Bernard GR, Dittus RS, Ely EW. Long-Term Cognitive Impairment after Critical Illness. N Engl J Med 2013; 369(14): 1306–16. doi: 10.1056/NEJMoa1301372.

22. Wilson RS, Hebert LE, Scherr PA, Dong X, Leurgens SE, Evans DA. Cognitive decline after hospitalization in a community population of older persons. Neurology 2012; 78(13): 950–6. doi: 10.1212/WNL.0b013e31824d5894.

23. Nikayin S, Rabiee A, Hashem MD, Huang M, Bienvenu OJ, Turnbull AE, Needham DM. Anxiety symptoms in survivors of critical illness: a systematic review and meta-analysis. Gen Hosp Psychiatry 2016; 43: 23–9. doi: 10.1016/j.genhosppsych.2016.08.005.

24. Parker AM, Sricharoenchai T, Raparla S, Schneck KW, Bienvenu OJ, Needham DM. Posttraumatic stress disorder in critical illness survivors: a metaanalysis. Crit Care Med 2015; 43(5): 1121–9. doi: 10.1097/ccm.0000000000000882.

25. Rabiee A, Nikayin S, Hashem MD, Huang M, Dinglas VD, Bienvenu OJ, Turnbull AE, Needham DM. Depressive Symptoms After Critical Illness: A Systematic Review and Meta-Analysis. Crit Care Med 2016; 44(9): 1744–53. doi: 10.1097/ccm.0000000000001811.

26. Amoretti MC, Lalumera E. COVID-19 as the underlying cause of death: disentangling facts and values. History and Philosophy of the Life Sciences 2021; 43(1): 4. doi: 10.1007/s40656-020-00355-6.

27. Mansfield KE, Mathur R, Tazare J, Henderson AD, Mulick AR, Carreira H, Matthews AA, Bidulka P, Gayle A, Forbes H, Cook S, Wong AYS, Strongman H, Wing K, Warren-Gash C, Cadogan SL, Smeeth L, Hayes JF, Quint JK, McKee M, Langan SM. Indirect acute effects of the COVID-19 pandemic on physical and mental health in the UK: a population-based study. The Lancet Digital Health 2021; 3(4): e217–e30. doi: 10.1016/S2589-7500(21)00017-0.

28. NHS Digital. BETA – Data Security Standards. 2020. https://digital.nhs.uk/about-nhs-digital/our-work/nhs-digital-data-and-technology-standards/framework/beta---data-security-standards (accessed 6th August 2020).

29. NHS Digital. Data Security and Protection Toolkit. 2020. https://digital.nhs.uk/data-and-information/looking-after-information/data-security-and-information-governance/data-security-and-protection-toolkit (accessed 6th August 2020).

30. NHS Digital. ISB1523: Anonymisation Standard for Publishing Health and Social Care Data. 2020. https://digital.nhs.uk/data-and-information/information-standards/information-standards-and-data-collections-including-extractions/publications-and-notifications/standards-and-collections/isb1523-anonymisation-standard-for-publishing-health-and-social-care-data (accessed 6th August 2020).

31. Secretary of State for Health - UK Government. Coronavirus (COVID-19): notification to organisations to share information. 2020. https://www.gov.uk/government/publications/coronavirus-covid-19-notification-of-data-controllers-to-share-information (accessed 6th August 2020).

